# “At the end of the day, it is Council’s decision”: Integration of health and equity into urban design and urban planning decisions and policies in Regina Saskatchewan

**DOI:** 10.1101/2023.12.05.23299446

**Authors:** Akram Mahani, Joonsoo Sean Lyeo, Agnes Fung, Kelly Husack, Nazeem Muhajarine, Tania Diener, Chelsea Brown

**Author notes:** **Corresponding author: Joonsoo Sean Lyeo**. **CRediT authorship contribution statement: Akram Mahani**: Funding Acquisition, Conceptualization, Data Collection, Methodology, Project Management, Writing, Review and Editing, Supervision. **Joonsoo Sean Lyeo:** Data Analysis, Literature Review, Writing the First Draft of Manuscript, Editing and Revising Different Drafts of Manuscript. **Agnes Fung:** Data Analysis, Preparing Tables & Figure, Contribution to Manuscript Writing. **Kelly Husack, Nazeem Muhajarine, Tania Diener, Chelsea Brown:** Conceptualization, Reviewing and Contextualizing Research Findings, Reviewing and Commenting on Different Drafts of Manuscript. **Declaration of competing interest:** The authors declare that they have no known competing financial interests or personal relationships that could have appeared to influence the work reported in this paper. **Data availability:** The data that has been used is confidential.

## Abstract

While there is a wealth of literature on the impact of urban design on health, our understanding of the factors that influence integration of health into urban design is limited. With the growing recognition of cities playing a leading role in enhancing health equity and population health outcomes, there is a need to examine the perspectives and experiences of municipal actors around health and equity. To address this gap, we interviewed 30 stakeholders engaged with urban design policy- and decision-making at the City of Regina in Saskatchewan, Canada. We found a lack of shared understanding of health among municipal actors. Our findings identified a number of factors that serve as facilitators and barriers to integrating health and equity in urban design policies. Findings from this case study deepen our understanding of these factors and provide recommendations for developing healthy urban design policies. Our findings underscore the importance of adopting an integrated and holistic approach for healthy and equitable urban design. As urbanisation continues to bring a greater share of the world’s population into urban areas, it is imperative that we deepen our understanding of how municipal governance can be leveraged to create environments that are conducive to the wellbeing of their residents.

## Introduction

The link between health and urban design and planning is not a new concept, as urban planning helped control infectious disease outbreaks in the 19^th^ century through sanitation (e.g., wastewater management systems, sewerage systems ^1^), and segregated zoning (e.g., separating polluting industrial development from residential areas ^2,3^), among other things.^1,4^ Leading international bodies such as the World Health Organization (WHO) have long recognized the impact of urban design on population health; for example, WHO suggests “placing health and health equity at the heart of [urban] governance and planning”.^5^ The United Nations (UN) Sustainable Development Goal 11 seeks to make cities inclusive, resilient, safe, and sustainable.^6^

The literature on the impact of urban design on health and equity is fast growing.^1,7,8^ Urban design has the potential to improve physical health (e.g., reducing non-communicable diseases^9^), mental health (e.g., depression^10^), and social health (e.g., high quality open spaces fosters social connection and cohesion^7,11^). There is also a growing body of evidence on the impact of urban design on equity due to the growing recognition of the social determinants of health (SDOH) – a term referring to the socioeconomic variables that shape an individual’s working and living conditions.^12^ SDOH are not evenly distributed throughout a population and contribute significantly to disease and premature death.^13^ As a result, some members of the population experience socioeconomic conditions that are either more or less conducive to good health.^14^ The unequal distribution of these conditions is the product of inequitable social policies, unfair economic arrangements, and underlying systems of oppression.^15^

There exists a complex and intertwined web of relationship between built environment and the SDOH, health, and equity.^13^ The built environment refers to “the physical characteristics of the areas in which people live including buildings, streets, open spaces, and infrastructure”.^16^ Each facet of the built environment is a purposeful decision that can either enable or inhibit an individual’s health.^7^ Examples of such decisions include: the layout of street networks and cycling infrastructure (e.g., physical activity^1,8^, risk of traffic accidents^17^, pollution exposure^18^, perception of safety and belonging^19,20^); the spatial distribution of grocery stores (e.g., access to healthy foods^21^); the choice of construction materials and dwelling design (e.g., energy use, light, fresh air, etc.^19,22^); and public building distribution (e.g., costs of accessing employment, goods and services, education, etc.^1,19^). In terms of equity, evidence suggests more health enhancing urban areas are disproportionately located in higher socio-economic regions.^19^ Despite this growing and strong evidence for change, effective strategies to influence integration of health and equity into urban design and built environment remain under-researched and underdeveloped.^23–25^

The rapid urbanisation globally – with Canada being 80% urbanized^26–28^ – has positioned cities and urban centres at the frontier of a number of global health challenges including climate change, environmental degradation, pandemics, and food insecurity.^1^ Addressing these challenges requires close collaboration between public health professionals and urban design professionals.^3,29^ It also requires community engagement and nonpartisan political leadership.^1^ Canadian cities are thus well-positioned to safeguard population health and reduce health inequities. By recognizing that urban design is intertwined with health, there is an opportunity to address the gaps that have historically contributed to the prevalence of longstanding health inequities.^30^ However, not all urban design professionals readily accept the responsibility for residents’ health.^31,32^ There are urban planners who do not consider health as part of their work.^31,33^ A study in Vancouver, Canada showed that municipal actors had limited knowledge of their role in reducing health inequities.^34^ In Canada, the socio-political climate (e.g., privatizing public programs to support vulnerable populations) has left municipalities with limited options to address public health challenges including health inequities.^34–36^ This disconnect between urban design and public health, has manifested itself in urban design around private automobiles and expansion of urban sprawl in Canada.^37,38^

Therefore, active transportation infrastructure such as bike lanes or wide sidewalks are not viewed as essential for community health, but rather a development cost.^39^ This has led to growing calls for inter-disciplinary collaboration between urban planners and public health professionals.^40,41^

In this paper, we present a case study of a medium-sized city in Saskatchewan, Canada detailing understanding of key municipal stakeholders of the link between health and urban design. Using the City of Regina as a case study, this study aims to investigate how health and equity are integrated into the urban design policies and decisions, in particular the opportunities and barriers to this integration. There is a bias in existing policy research to focus on the experiences of large metropolitan cities rather than the small-to medium-sized cities that are representative of a larger share of cities worldwide.^42^ As a result, many of the recommendations emerging from this body of research have been developed in considerations of the needs and capacities of larger cities, meaning that they must be scaled down to the contexts of medium-sized cities.^42^

While there is a wealth of literature on the impact of urban design on health, our understanding of the factors that influence integration of health into urban design policies is limited. Findings from this case study deepen our understanding of these factors and provide recommendations for developing healthy urban design policies. Given the role of municipalities in enhancing population health outcomes, there is a need to examine the perspectives, beliefs, and experiences of municipal actors who have been engaged with policy- and decision-making around urban design and built environment.

### Conceptual Framework

To better conceptualize the link between health and built environment, we adopted the Healthy Built Environment Framework developed in 2018 by the BC Centre for Disease Control.^43^ This framework identifies five key elements of a healthy built environment and pathways (causal linkages) between urban design and health (see Figure 1) including:

1. Neighborhood Design: Healthy neighborhood design is facilitated by land use decisions which prioritize complete, compact, and connected communities.
2. Transportation Networks: Healthy transportation networks prioritize and support active transportation modalities.
3. Housing: The design, quality, and affordability of diverse housing options has a critical influence on health and well-being.
4. Food Systems: Accessibility and affordability of healthy foods can be supported through land use planning and design.
5. Natural Environments: Community planning which preserves and connects the surrounding natural environment can have significant health and well-being impacts (green space).

**Fig 1.**
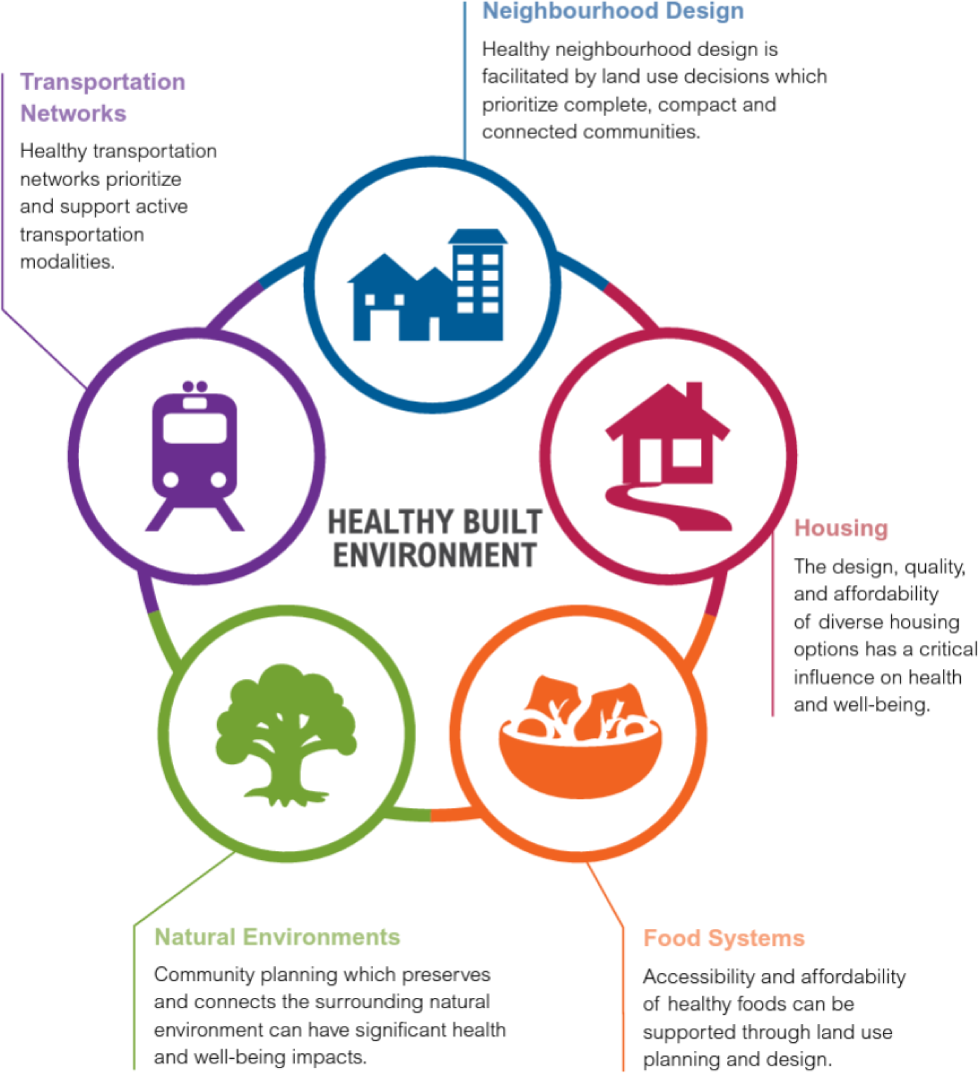
The Healthy Built Environment Framework ^43^

## Materials and Methods

### Research Setting

The City of Regina is the capital and second-largest city of the province of Saskatchewan. With an estimated 226,404 residents, Regina is home to roughly 20% of the province’s population and is the twenty-fourth largest municipality in all of Canada.^44^ While a medium-sized city, the City of Regina has seen steady growth over the past few decades – a culmination of natural population growth, a provincewide trend in rural-to-urban migration, and the growth of international immigration to the prairies. ^45,46^ By 2040, the City of Regina is projected to reach a population of 300,000.^47^ Today, immigrants and Indigenous peoples account for approximately 20.3% and 10.4% of the city’s residents.^44^ This can be contrasted from two decades prior, in 2001, when these groups respectively comprised 7.4% and 8.7% of the population. In recognition of these demographic shifts, decision-makers in the City of Regina have sought to understand how the city’s physical form and built environment must be adapted considering these changes.^47^

The municipality of Regina is governed by the Regina City Council, which consists of eleven elected representatives, one Mayor and ten City Councillors,^48^ who are elected every four years – the Mayor being elected by voters from the City of Regina at-large, while City Councillors are elected by voters from the specific wards they represent. In addition to their decision-making role, City Council is responsible for appointing the City Manager, who is directly accountable to the City Council and tasked with overseeing the four divisions of the City’s operations: (1) Financial Strategy & Sustainability, (2) City Planning & Community Development, (3) Citizen Services, and (4) the Transformation Office.^49^ The City Manager also provides strategic advice to City Council as an Officer of the Council, a role they occupy alongside the City Clerk and the City Solicitor. The City Manager and the divisions they oversee collectively make up the City Administration. The City Administration is responsible for executing the day-to-day operations of the municipal government.

In this study, ‘decision-makers’ refer to elected officials responsible for choosing policy directions and priorities, deciding which policies will be translated into tangible action by the municipal government.^50^ This responsibility chiefly falls onto the City Council and the Mayor. The term ‘policy-makers’ refer to public servants responsible for developing policies in accordance with the mandate chosen by decision-makers.^50^ In Regina, this work falls onto the City Administration. While policy-makers can make recommendations and submit proposals, they often have comparatively little influence over the overall direction of policy relative to decision-makers. The term ‘stakeholder’ is used in reference to both policy-makers and decision-makers.

### Data Collection

The first author (AM), who is a trained qualitative researcher, conducted all interviews (n=30) between May and July 2023 using the video conferencing platform, Zoom. With participants’ consent all interviews were audio-recorded and transcribed verbatim using Otter.ai, a speech-to-text software. Each interview lasted a minimum of 60 minutes. An interview guide/protocol was developed through research team expertise and engagement with existing literature (see Appendix 1). Some demographic information such as education background, years of experience in the profession and in their current position were obtained at the beginning of each interview. All illustrative quotes were checked by research participants (member checking).

### Sampling and Recruitment

We used a combination of purposive, snowball, and convenience sampling strategies to recruit our research participants. Guided by the Healthy Built Environment framework^37^ and SDOH^12^, we purposefully recruited our research participants from the City Administration divisions including: (1) Financial Strategy & Sustainability, (2) City Planning & Community Development, (3) Citizen Services, and (4) the Transformation Office. We also included City Councillors to have decision makers perspectives. The City assisted with distributing our recruitment letter among divisions listed above and those interested (n=4) contacted the first author directly (convenience). One of the Research Advisory Board is a member of City staff who also assisted with identifying potential participants. We also collected a list of City Councillors from the City website. Participants were eligible if they were a City staff who had been engaged with policy- and decision-making around urban design and urban policies related to SDOH and any of the key elements of the Healthy Built Environment Framework. Potential participants were also identified through snowball sampling through their colleagues.

### Data Analysis

The interview transcripts were analysed using a qualitative thematic framework.^51^ Using this approach, transcripts were investigated for recurring patterns in how the interviewees responded to the questions asked.^52^ The differences and commonalities between these recurring patterns were used to categorise their responses into distinct themes, with each theme grouping together interview transcripts that communicated a shared implicit or explicit message. When appropriate, themes representing broad ideas were broken up into smaller sub-themes focusing on specific elements of said higher-level ideas. Themes and sub-themes acted as the basic unit of our analysis, and ultimately informed the explanatory conclusions that were drawn.

This analysis employed a mix of inductive and deductive analytical methods.^53^ Our initial data analysis was informed by nine key themes developed *a priori* based on interview questions (see Table 2). The interview transcripts were first deductively analysed, a process in which transcript excerpts were actively organised into these existing themes. We then employed a constant comparative methodology inherent to a grounded theory approach to, when appropriate, establish new emergent themes *a posteriori*.^51^ This approach served as the basis for our inductive analysis, which involved a ‘reflexive’ and flexible approach to our analysis.^54^ Continuous dialogues and discussions within our multidisciplinary research team, that included knowledge users from both the public health system and the municipal government, encouraged and enhanced reflexivity, trustworthiness, and credibility.^54^ Throughout the analysis, regular peer debriefing took place to explore whether the initial findings correspond with their expertise.^55^ Each team member looked at the data from a unique and different lens, but through dialogues, discussions, and constant comparisons with the data we ensured our analysis is grounded in the data. This analytical process was completed by two members of research team (JSL, AF), who each reviewed the interview transcripts to organise relevant excerpts into corresponding themes and sub-themes. This was an iterative process, in which the team members frequently met to resolve conflicts and to review, revise, and refine the emergent themes and sub-themes. We used NVIVO 12 qualitative data analysis software to support the thematic analysis and subsequent identification of relevant themes and sub-themes.

**Table 1.**
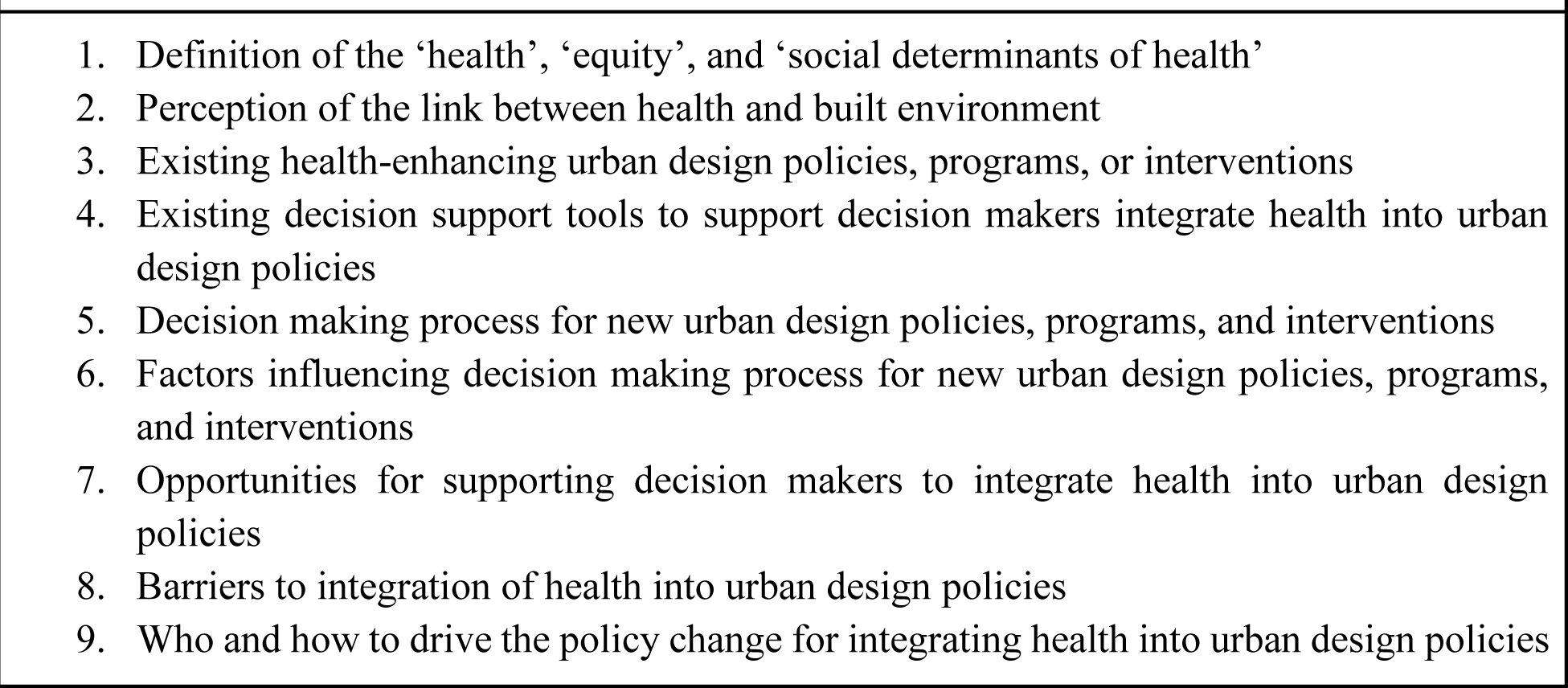
Qualitative themes identified a priori for deductive analysis.

**Table 2.**
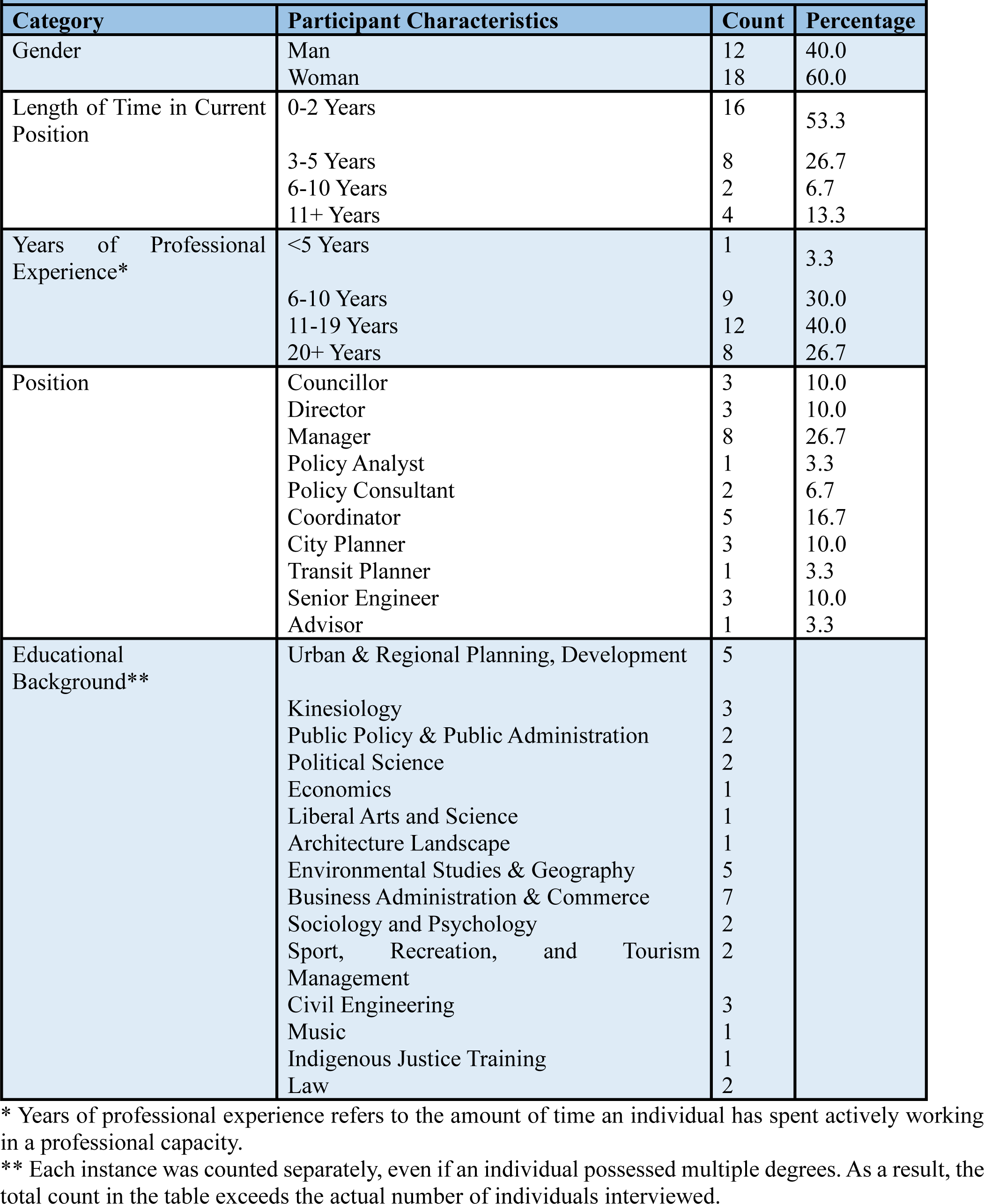
Characteristics of interviewees (n = 30)

### Ethical Considerations

Ethics approval for this research was received from the University of Regina Research Ethics Board (project ID: 2023-109). All research participants received verbal and written information about the objectives and voluntary nature of the study. Written and verbal consent were obtained from all interviewees. The interviewer removed all identifying information from the transcripts.

## Results

### Characteristics of Participants

Sixty-five potential participants were contacted, of which 30 agreed to participate in the study, a response rate of 46%. Research participants had different role/status (e.g., Councillor, Director, Manager, Coordinator, Advisor) and diverse education backgrounds. Most participants (n=17) were from the City Planning & Community Development division given the direct role of this division in urban planning and urban design. Participant characteristics are summarized in Table 2.

### Understanding of Health and Equity

The way our interviewees perceived, defined, and understood the concepts of health and equity often influenced how these ideas guided their development of urban planning and urban design policies and decisions. Interviewees shared examples of how gaps in the understanding of health and equity, both within themselves and among their peers, acted as a significant barrier to meaningful action in policy- and decision-making Two key themes emerged around how our participants understood health and equity including: paying lip service to equity; and lack of a shared understanding of health.

### Paying lip service to equity

There was a common belief among our research participants that equity is sometimes integrated into policy only at a surface level. All interviewees acknowledged the importance of equity in their work; however not all interviewees believed that they could sufficiently integrate equity into their policy or decision-making.

One major barrier to the integration of equity by the City Administration was the lack of a shared understanding of what equity means. Our research participants had differing interpretations of equity, resulting in differing or even contradictory approaches for how equity could be pursued. The lack of a standard definition of equity, or even a common language to facilitate dialogue around equity, was described by one participant as being a source of conflict and friction:

> *“And I think we can all sit in an office and decide what we think is equitable, or what we think is equal, and what we think our definitions of them are, but every human is going to have their own definition of what is equitable or equal to them. And so I’ve actually recently kind of started to not like having even the conversation of equitable versus equal. And instead, I like to more talk about what are the choices.” [P20].*

Another participant noted:

> *“The way I would characterise the dominant model of equity is everybody at the City would be aghast if they found out a particular person was without a house, because they were a queer person. But nobody is upset that there are a lot of queer people without houses for some other reasons. And so the principle isn’t queer people should have houses or everybody should have a house, it’s nobody should lack a house simply because they’re queer. But if they just happen to lack a house and be queer, well, so be it. And I think that highlights differences in our definitions of equity, whereas I would say it’s about having this base model, which includes a house, they would say, well, as long as you’re not out of a house because of discrimination so be it.” [P26]*

Furthermore, many interviewees observed that while they considered the integration of equity to be important to their work, this view was not necessarily held by all members of the City Administration. For instance, there were several instances where interviewees described interactions with peers who were dismissive of the integration of equity into urban planning decisions. One interviewee described how their peers waived concerns about equity under the pretence that such concerns were unnecessary:

> *“There are some people I think, who have this idea, they say: ‘well, I don’t see colour, or gender, or whatever, I just support the best person for the job’; but they don’t look at, okay, there’s systemic structural issues that are maybe creating racism.” [P25]*

Several interviewees also expressed their concern that efforts to integrate equity into municipal action were largely limited to token efforts. In such cases, policies were criticised for giving the appearance that equity was being pursued, without meaningfully supporting the integration of equity. There was an argument that some decision-makers have come to recognize that the concept of equity has gained traction with large swathes of the public. In light of this, some decision-makers have pushed forward policies with equity-based messaging to attract or appease voters, without necessarily ensuring that these policies are having any tangible impact in practice. As stated by one interviewee:

> *“It does feel at times, like we’re putting forward that equity term, just to check the box, right? That’s when we run into the problems when it comes to, okay, what are we doing tactically to move this forward? Well, now we don’t know, because we haven’t really defined that well in the first place, right? And so what does that mean? That’s always a question.” [P13]*

Interviewees suggested that stronger leadership is needed to overcome these barriers. Participants argued that this responsibility ultimately falls on the decision-makers including the Mayor and City Council. Because decision-makers are typically seen as authoritative figures within the municipal government, they are well-positioned to provide clear and consistent messaging on what the integration of equity into urban planning should look like. They have the opportunity to outline specific objectives and benchmarks to guide policy-makers in the incorporation of equity into their work. One interviewee described the need for leadership as follows:

> *“…if leadership was prioritising equity and health and committing the resources in order to provide, like staff training and staff positions, like to be able to kind of keep the work moving and get people who are more reluctant on board, I think that would help. I think, yeah, and then creating, like just fostering a culture throughout the corporation of this needing to be a priority. And so I think, like in some ways that would need to come from Council and executive leadership.” [P7]*

### Lack of a shared understanding of health

We observed that there were some discrepancies in how interviewees understood the concept of health. The term wellbeing was more frequently used by our interviewees compared to health or wellness. There was a lack of consensus among interviewees on the extent to which certain traits were perceived as being integral to an individual’s health. When interviewees were asked to define health, there were some traits that were almost always mentioned such as physical dimension, and other traits that were mentioned more rarely such as social dimension. When describing physical health, interviewees often referred to characteristics that could either be visually identified (e.g. the presence of an injury) or quantifiably measured (e.g. blood pressure) as one interviewee stated:

> *“First thing that comes to my mind is really just the physical wellbeing of a person, obviously, so any type of ailments and whatever else would contribute to poor health, but then obviously, physical characteristics like, well it’s hard to describe health, because it’s like, well, how healthy are your lungs? Or do you have good blood pressure? Or do you have the proper vitamins, minerals…” [P18]*

In the few instances where the City Administration had the opportunity to engage with the public health professionals (i.e., Saskatchewan Health Authority), conversations around health were mainly limited to projects that interpreted health through the lens of physical health (e.g., encouraging exercise and physical fitness).

It was also fairly common for participants to consider mental health an important and integral dimension of health. This widespread consideration seems to be a relatively recent development reflecting broader societal shifts in how health is understood in the public consciousness as one participant noted:

> *“I think now there’s been more of a focus on mental health. But, I think probably growing up, and probably till five years ago, it was really about physical health. So, you know, exercise, diet, nutrition, all that kind of stuff. But there’s probably been a real focus now, especially with the pandemic, and that kind of stuff on more mental health and mental wellbeing.” [P5]*

In contrast, fewer interviewees extended their definitions of health to include a social dimension – at least, not before being explicitly prompted with a question asking them to define social determinants of health.

> *“Health is more complicated than just, I feel, I think typically people think of physical health, but I mean, I very much appreciate that health is really multifaceted. [It] involves physical, mental, spiritual. Our health and our mental health can be impacted by all sorts of things and communities from recreation to social inclusion to…” [P28]*

A few interviewees also used the WHO (1948) definition of health. Participants cited examples of how health was often interpreted to mean different things to different stakeholders as one interviewee noted:

> *“I’m thinking about our Council members and the decision-makers. And I think the struggle with that is an understanding of what health is, because what health is to one Councillor may be different than what it is to another. So, I think what we don’t have at the city is a good collaborative understanding of what health is.” [P6]*

The current lack of a consistent and shared understanding of health among stakeholders was described as a significant barrier to meaningful action, and integration of health into urban planning as opportunities for integration are limited by a lack of shared motivation, knowledge, or interest among key stakeholders.

> *“I don’t think our [design] standards have ever really been looked at from that health perspective. And I think a lot of what drives from the developer side is the market and what people are willing to buy.” [P5]*

Our interviewees described the current situation as a reactive, rather than proactive, approach of the municipal government to health as one participant noted:

> *“One of the things that I noticed as a professional in this area is that health promotion is not really discussed. It’s discussing a lot of the issues that they’re dealing with currently at this moment. So it’s got a great foundation, a great platform, great start. But we don’t talk about holistic, how do we promote health so that we don’t have to get to this point where we’re addressing issues.” [P6]*

### Opportunities to Integrate Health in Urban Design

Interviewees described several key events that facilitated the integration of health into urban design. These events were often associated with the opening of policy windows – sweeping changes in broader public sentiment, or at least the sentiment of key stakeholders – that resulted in conditions that were favourable to the passing of relevant policies. These opportunities include: giving a voice to disenfranchised communities; the COVID-19 pandemic opening a policy window; and the role of champions and leadership in policy change.

### Giving a voice to disenfranchised communities

Several interviewees acknowledged that, within the City of Regina, there are certain sub-populations that have struggled to advocate for themselves and engage in the policy- and decision-making process. Oftentimes, this is the result of historic and ongoing inequities that have culminated in the disenfranchisement and marginalisation of certain communities. For instance, the City of Regina has a sizable Indigenous population – comprising more than 10% of the population – that has long been underrepresented in leadership roles within the municipal government. One interviewee described how efforts to elevate and empower Indigenous voices have presented the City of Regina with new opportunities that would have been missed otherwise.

> *“When looking at Indigenous populations in Regina, and the nature in which, like narratives and history are told within that, I think that might help sort of push that change to a more qualitative look at things. Partially, it’s a competitive advantage that Regina might have, that other municipalities don’t.” [P9]*

Interviewees also acknowledged that the empowerment of disenfranchised communities is not just a matter of strategic advantage, but also a matter of justice and fairness. In many instances, vulnerable populations are not only less likely to have the social and political capital to advocate for themselves, but are also disproportionately more likely to be overlooked – or even negatively impacted – by policy developments. One interviewee described how members of disenfranchised communities have been given an active voice in recent policy- and decision-making initiatives, allowing for the development of policies that meet their needs.

> *“Part of it is trying to ensure that the people who are going to be most impacted by our decisions are consulted, if not hired… So that people who are making decisions about our recreation program or how we communicate are actually informed by their own lived experience of disability or marginalisation in some way, and have those connections with community members.” [P7]*

Lastly, some interviewees tied the importance of empowering disenfranchised communities to the democratisation of the decision-making process. These interviewees noted that decision-makers, as elected officials, are often hesitant to support policies that do not appear to have a substantial public backing. Addressing barriers to participation that many disenfranchised communities face is instrumental to ensuring proportional representation and consideration in the policy- and decision-making process. Interviewees perceived the role of city staff to educate and inform the public/community in their engagement activities.

> *“Staff can provide good recommendations. However, if Councillors don’t feel that interest and push from the community, it’s hard for some of them to make that leap. For them it may feel risky. One thing that I find interesting over the last few years is the people who come to delegate to Council – it’s increasingly diverse, it’s not just the same few people who come to every meeting. You’re starting to hear from people who don’t normally come to meetings and share their perspectives.” [P30]*

### The COVID-19 pandemic opening a policy window

The COVID-19 pandemic resulted in significant disruption to the status quo, dramatically altering how health – especially public health – is positioned in the public consciousness. The onset of the pandemic saw a widespread systems-level shock pushing public health messaging to the forefront of public attention, which led to a widespread shift in the way many individuals, both in government and in the wider public, understood the importance of health. Similarly, it seems that the COVID-19 pandemic, especially during the height of lockdowns, resulted in a shift in the way people engaged with their physical surroundings. With indoor venues being shuttered and perceived as unsafe, public outdoors spaces – such as gardens, parks, and urban green spaces – saw increased traffic as places for recreation, socialisation, and health-enhancing activities. The City has since continued to see increased traffic to outdoor public spaces, as well as calls to expand the number of outdoor spaces available to the public. This sentiment was captured by the following quote:

> *“I think one of the things that COVID really elevated is the increase in importance of our public spaces. So I know that parks and outdoor facilities are busier than they have ever been before. Because throughout the pandemic, people were using these public spaces that they hadn’t used before. And now they value it. And so they’re asking for more.” [P6]*

The COVID-19 pandemic thus opened a policy window in which policies that targeted health, and public outdoor spaces were more likely to be seen favourably. Several interviewees remarked that since the onset of the COVID-19 pandemic, there has been a push by key stakeholders within the municipal government to allocate resources and political willpower to the expansion and maintenance of public outdoor spaces. One interviewee stated:

> *“We built about 1.6 kilometres of [multi-use] pathways during the COVID pandemic, and that was all the funding from the provincial and federal governments, the Municipal Economic Enhancement program, I think. So these big programs, they’re more common, this isn’t a thing that, like we were seeing three or four years ago.” [P9]*

### Role of champions and leadership in policy change

Leadership was widely recognized as a major facilitator of meaningful policy change. Interviewees explained that they often looked to the leadership of *champions* in the municipal government. The term champion was often used in reference to highly-motivated individuals – often in positions of authority – who were able to dictate the course of policy action. These champions often had clear and actionable goals, which they would use to coordinate the actions of their teams. The direction provided by champions could facilitate the mobilisation of large amounts of resources – e.g. capital, manpower, political willpower – in pursuit of specific objectives.

Several interviewees pointed to the Mayor as an example of a champion who leveraged their position in the municipal government to prioritise certain policy objectives on the municipal agenda. At the time of data collection, the Mayor was regarded as a champion who was able to bring conversations on the social determinants of health to the forefront of municipal decision-making. The Mayor was seen as someone who could coordinate diverse stakeholders to redirect their attention in pursuit of specific goals.

> *“I just want to comment on the Mayor, and I have to say that, well, I have only been with the City for [X] years. But it was the first time that I really saw a leader, gather people from the community and government, and community organisations together in rooms to talk about specific problems, like domestic violence and intimate partner violence, and homelessness, and all sorts of things. And she did a fantastic job, getting those conversations started, and getting people talking about them. And let’s work together on these.” [P17]*

### Systemic Barriers to the Integration of Health into Urban Design

Interviewees identified several systemic barriers to the integration of health in urban design policies including: inaccessibility of evidence; insufficient resourcing; fragmented governance structure; limited legal power of local governments in Canada; and an entrenched culture of individualism and libertarian ideology. These barriers were often entrenched and perpetuated by institutions, systems of government, or wider beliefs – and generally could not be overcome through the efforts of an individual stakeholder acting alone. These barriers made certain policy actions infeasible or impractical in the status quo.

### Inaccessibility of evidence

Evidence was described as an essential building block of policy- and decision-making. Interviewees often acknowledged the value of informing policies and decisions by evidence. However, in practice, many interviewees explained that they faced significant barriers to accessing evidence preventing their policies being informed by the best possible evidence available. For example, one interviewee described how administrative barriers, namely a lack of coordination between the provincial and municipal governments, posed a barrier to the uptake of evidence in policy-making:

> *“We were seeking data from the province on the number of licensed facilities, and they weren’t very forthcoming with even that kind of basic information. So to me, if we want to get into more nuanced data, if we want to apply that health lens, like in a meaningful way, we really do need adequate data to back that up. And I don’t think we’re there yet in terms of relationship with the province or their willingness to provide that.” [P13]*

Other interviewees pointed to barriers in accessing the scientific evidence produced by academic community. In some cases, these barriers were associated with a prohibitive financial cost – for instance, when access to scientific studies is locked behind a paywall. In other cases, the scientific evidence produced by academia was described as being created in a ‘black box’. Several interviewees described instances where their ability to read, interpret, or understand academic studies was hindered by the use of jargons and technical language as one participant noted:

> *“There’s a lot of academic acumen that’s used and terminology, and it can be overwhelming, and nobody wants to walk out of a room and feel stupid.” [P6]*

Sometimes, the value of using data to inform decisions and policies is not well understood by the government. To address this, interviewees highlighted the importance of knowledge translation and translating research recommendations to the goals of policy- and decision-makers in an accessible way.

> *“I think there is more that academics could do to help policy-makers understand how they reach their conclusions. I think they [academics] have legitimate claims to expertise, and why areas within their expertise ought to be given more weight.” [P26]*

> *“…we need individuals that can distil that [academic] information, and provide it in a more palatable, and more understandable format.” [P6]*

### Insufficient resourcing

Many interviewees described instances where they felt limited in their ability to enact change due to a lack of resources. These resources could take different forms. One of the most common resource limitations discussed by interviewees was a lack of funds. In Canada, these financial constraints are exacerbated by the limited revenue streams available to municipalities. With the exception of transfer payments from the federal or provincial governments, which are often tied to specifications for how the money must be spent, municipalities are largely reliant on property taxes to finance the bulk of local services and expenditures. As a result, municipalities often have limited flexibility to levy funds for municipal projects. This was recognized by one interviewee, who stated:

> *“They’re being asked to do more with the same. I think it all comes down to resource allocation. We put our money where we are individually, and we put our money where we think things are important to us, where our values are, and the values are not necessarily aligning with the allocation.” [P6]*

Other frequently mentioned resource limitations include a lack of time and staffing. However, these limitations are often directly related to funding. For instance, while many interviewees expressed that they faced time pressures to develop policies, these time pressures were often related to the inability to fund policy projects indefinitely. Similarly, while many interviewees expressed that their teams often lacked the operational capacity to implement policies to their full extent, these constraints were often related to the lack of funds needed to hire more staff. One interview discussed the interrelated nature of these resource limitations as follows:

> *“Change does not happen overnight, it takes a lot of time, sometimes it takes a cultural shift within City Hall. Sometimes it takes new positions to be allocated, people specifically devoted to advancing whatever goals there are. And a lot of times, it’s just not easy to hire staff, cities get into budget crunches, and decisions are made to collapse portfolios, and combine them into others and then things kind of get watered down or kind of lost, or they start to slow down.” [P27]*

### Siloed or fragmented governance structure

Siloed or fragmented governance structure and lack of an integrated approach to urban design was identified as a significant barrier to meaningful collaboration and coordination between policy-makers from different branches and divisions of the municipality. This lack of coordination is often referred to as ‘siloing’, wherein stakeholders are so locked into their own respective spaces that they are unable to capitalise on the resources from those in other spaces. This results in missed opportunities for a comprehensive and multi-disciplinary collaboration to promote healthy urban design. This siloing can lead to the emergence of differing institutional logics, epistemologies, and cultures within and across silos. These differences often present a significant barrier to cross-sectoral work across silos, making it difficult to share knowledge and collaborate on projects. For instance, one interviewee explained how differences in the use of jargon could create confusion, impeding cross-sectoral collaboration.

> *“Often it’s just language we’re using. So when I was working with architects, they used the word program very differently than I do as a [X] planner. And I didn’t realise that until a few weeks in and I went, Oh, you’re using a program as a different term.” [P6]*

Interviewees also described instances that siloing could lead to conflicts between objectives of different branches and divisions. One such example was the opposing objectives of active transportation planners, urban designers and landscape architects, and traffic engineers. An interviewee stated that while these professionals tend to be more concerned with the movement of people, traffic engineers are more concerned about the movement of vehicles. This discrepancy, while subtle, resulted in conflicts as described by the interviewee:

> *“Often, the natural adversary of good urban design and opportunities for active transportation is, or has been, the traffic engineer. There were lots of policies that we seem to put in place that very much favour the movement of vehicles over the movement of pedestrians, cyclists”. [P1]*

### Limited legal power of local governments in Canada

In Canada, municipalities are widely considered to be ‘creatures of the province’ or ‘a third order of government’, which refer to the differences in the legal powers of municipalities relative to provinces. While provinces derive their legislative powers from the Constitution – and thus have provisions guaranteeing the permanence of their legal powers – municipalities instead derive their legislative powers from provincial law. In practice, this means municipalities are generally only able to exercise powers that have been delegated to them by the provincial government. Furthermore, provincial governments have the ability to modify or revoke the powers delegated to municipalities at any time, as long as they can gain the necessary share of votes in the provincial legislature required. One interviewee outlined the consequences of the municipal-provincial relationship as follows:

> *“The cities are an entity of the province, the province is not an entity of the federal government, right? The relationship is different between cities and the province, and so the province can really impact some of our policy choices, and really impact whether we have the money to fund things, they’re the ones who set what’s required around municipal reserves.” [P16]*

The ack of clarity around the roles and responsibilities of municipalities and provincial government was reported as a source of tension, which is costing the municipality as the following two quotes demonstrate.

> *“The Ministry of Education does not fund playgrounds. So they will build a school, but they will not build a playground. And so if your school wants a playground, you have to fundraise for that. There are things that are very much provincially driven that impact municipal health because again if you think about the mental and physical wellbeing of children, a playground is pretty key.” [P16]*

> *“There’s disagreement with how we interact with the province and we’ve kind of gone through a whole cycle in the last two to three years of whose responsibility is this? Like, does the city have a role, but then around some issues, we’re seeing that political narrative draw back to what it was before, because we’re just seeing that the Council’s assessing that the levying of property taxes for social issues is too much of a burden on property owners.” [P4]*

### Facing an entrenched culture of individualism and libertarian ideology

Some interviewees explained that they faced barriers due to the friction between collectivism – i.e. maximising benefits to the community as a whole and the prevailing attitudes of individualism in Canada. This can be seen when individuals are unwilling to sacrifice individual benefits – e.g. personal vehicles, lower taxes, etc. – in favour of policies that would benefit their communities as a whole. Attempts to integrate health into urban design are rooted in the idea of collectivism. Interviewees argued that the dominant individualistic ideology is reflected in the urban design, as seen in the prevalence of urban sprawl and road networks. As discussed by one interviewee, the prevalence of car ownership can be regarded as emblematic of how individualist cultural attitudes can influence the physical form of urban areas:

> *“North American transportation systems mirror that libertarian individualism, like the very existence of having a car is intrinsic to feelings of identity and liberty and the ability to sort of, like freely move at any one point. And there’s a lot of wrapped up sort of suppositions and considerations within that which are sort of strange, like I am entitled to drive anywhere, I’m always entitled to have my car, I’m always entitled to have a parking space, there is a dedicated on street parking space for my property in front of my house.” [P9]*

Many interviewees also noted that these individualist cultural attitudes often affected how policy issues were positioned in the public consciousness. Interviewees discussed how policy proposals were more likely to be met with resistance if they were perceived as infringing on individual liberties in favour of a collective good. One interviewee discussed the public backlash that accompanied a policy proposal to restrict the types of pesticides that an individual could use on their properties:

> *“With the pesticide thing, I mean, I think the PhD in biology was persuasive and explicit that pesticides do not want to stick to property lines, and will get onto your neighbour’s line. And it was thought, I think, in the debate: ‘That’s fine. That happens. I don’t need to consider the impact on my neighbours when I do these things, because this is my property’.” [P26]*

In line with the aforementioned friction with individualism, many policy-makers found it difficult to push forward policies that only a minority of people would benefit from. Stakeholders seemed less likely to support a policy proposal if they did not believe it would be personally advantageous to do so. Conversely, stakeholders also seemed more likely to oppose a policy proposal if they believed it would negatively affect them, even if they recognized that it would create a disproportionately greater benefit for another population. For instance, one interviewee described how policy proposals to establish safer conditions for sex workers were often met with fierce opposition, chiefly by individuals who did not want to see sex workers in their neighbourhoods:

> *“I feel like our Council right now is really grappling with the same sorts of polarised cultural issues that are impacting everybody widely. Here we are in Saskatchewan, which is quite conservative. For example, I think sex workers are mostly relegated to the industrial areas of the city, where there’s fewer police and fewer bus, public transit, and fewer lights. And there was just a conservative element on Council that was more interested in flowerpots around the city than in keeping sex workers safe.” [P17]*

### Navigating Decision-Making in Urban Design

There is no single, straightforward path wherein a policy is directly translated into the real world. The decision-making process is dynamic and subject to a wide selection of occasionally competing and occasionally collaborative forces. Many of the interviewees cited key nodes in the decision-making process that had significant influence on the way in which a policy is translated into the real world including: the City Council as the final arbiter of the decision-making process; tug-of-war between enacting change and maintaining the status quo; lack of representation; inseparability of decision-making from the political environment; and relationship with the private sector.

### City Council as the final arbiter of the decision-making process

The City Council is the main decision-making apparatus of the City of Regina. Before major policies can be put into action, they must first pass a vote from members of the City Council. This arrangement means that the elected representatives of the City Council, comprising the Mayor and the City Councillors, carry the most influence in the entire policy- to decision-making process. Many interviewees explicitly recognized the City Council as the final arbiter of the decision-making process – the bottleneck by which policy proposals were either incorporated into practice, or discarded:

> *“We as planners, our role is to do the research, find the data, make a recommendation, be there to support Council, and its discussion, but at the end of the day, it is Council’s decision.” [P27]*

To receive the assent of the City Council, interviewees acknowledged that their policy proposals had to be intentionally packaged in a way that appealed to a majority of seated members of the Council. For example, one interviewee explained that they found Councillors were more likely to be influenced by appeals to emotion through storytelling and personal accounts, especially when these appeals to emotion were perceived as coming from members of the public. In order to adjust their strategy accordingly, this interviewee pivoted their approach away from policy proposals that were contingent on the relaying on facts and statistics, and instead opted for an approach that was rooted in elevating the perspectives of members of the public:

> *“[In decision making process], they [City Councillors] are less swayed by narratives from us and far more swayed by residents. So, when a resident comes and tells us a story, a really personal story, or a really impactful story, I would say they’re very moved, and they can be moved by one story.” [P16]*

As stated by another interviewee, this arrangement was sometimes perceived as a source of frustration by policy-makers:

> *“More and more City Council tends to refer less to the City Administration as a base for the recommendations or decisions or to their committees like Regina Planning Commission. It used to be very predictable, that we provided advice to proceed in a particular direction, and then the City Council would more often accept it. And now there is much less predictability.” [P27]*

### Tug-of-war between enacting change and maintaining the status quo

It was widely acknowledged that nearly all policy proposals put forward by the City Administration had a corresponding, alternative option – the option of doing nothing and maintaining the status quo. Many interviewees referred to instances where key stakeholders, such as decision-makers and residents, were hesitant to support policies that would result in anything more than a minimal deviation from the status quo as the following quotes demonstrate:

> *“It’s easier, I think, to make new policies on something that’s brand new. And, people are more willing to accept that. But changing old policies that have been in place for decades is challenging. And me even trying to enforce or adhere to policies that are so outdated is certainly a challenge.” [P10]*

> *“…there’s a lot of resistance from people for changing their neighbourhoods, or changing the housing options in their neighbourhood. So that is a really difficult thing. And that I always hear people say, but we’re Regina, we’re not Montreal, or Calgary or Vancouver. So there is a limits in understanding that we can’t continue to expand and have the same kind of models, right? It’s just not sustainable. And it costs a lot for the City and to provide the services to a city if it continues to sprawl.” [P25]*

In some cases, interviewees explained that the general public’s preference for the status quo was so strong that perceived deviations from the status quo were met with significant backlash. Under these circumstances, the status quo was often positioned by decision-makers as the ‘path of least resistance’. This occurred even when decision-makers were presented with alternatives that they perceived to be better for the overall health of the general public. In other words, decision-makers were more willing to accept the maintenance of a lower quality – but politically palatable – status quo over a higher quality – but more politically contentious – alternative. This very same situation occurred when the City Council delayed approval for the fluoridation of Regina’s drinking water due to public outcry, even despite being presented with substantial evidence of the health effects of fluoridation:

> *“In the case of fluoridation, I think that was more: “Is it good? Yes. Is our current policy of not fluorinating harmful? No.” Therefore, if there is this crazy outcry of “No, we don’t want it in the city”, [the City Council] is willing to accept it because it’s not actively doing harm. It’s more just status quo, which isn’t necessarily a problem at this time. So they’re not willing to make a huge stink and issue about it.” [P18]*

Increasing polarization may also influence how relevant stakeholders perceive social issues that disproportionately affect marginalised populations under the status quo. In the context of this polarisation, there are concerns that sectarianism may foster an environment in which the majority ‘in group’ – which tends to enjoy the most privilege under the status quo – is unwilling to allocate time, resources, or political will to meet the needs of a minority ‘out group’. This may be especially pressing in cases where deviations from the status quo are perceived as some sort of loss to the majority in group.

### Lack of representation

Regina is a diverse city; however not all of its communities have been fairly represented in municipal policy- and decision-making. Interviewees acknowledged that, oftentimes, the populations that face the greatest barriers to participation also experience the most socioeconomic disadvantage. As a result, the populations that stand to benefit the most from being included in the development of new policies tend to be the least likely to have their voices taken into consideration.

Several interviewees acknowledged that, as a result of barriers to participation, only a minority of voices were allowed to influence the policy- and decision-making process. It was often recognized that this segment of the population represented a greater level of socioeconomic advantage than the City of Regina as a whole – comprising those who were able to leverage their social, economic, or political capital to have their ideas pushed to the forefront. It is important to consider why these voices, and the communities they represent, are better situated to exert their influence on the direction of policy action. In the political arena, the communities with greatest need for change may be the least empowered to fight for and advocate for themselves. Policy- and decision-makers must thus ensure that they are simultaneously responding to the needs of the public, while also not letting the loudest voices disproportionately dominate the conversation. One interviewee recognized how this arrangement may be, at least in part, shaped by the inaccessibility of the public consultation process:

> *“It’s tricky, because I feel like it’s almost the voices you don’t hear from a public consultation perspective. I’m always thinking about, well you have a [public consultation] session and you have 15 people show up. Well, what about the other 1000 that didn’t come out, right? Is it that this isn’t a concern? Is it that the session wasn’t accessible? Sometimes I think that people have kids and lives and all that kind of stuff. Or is it just people, I don’t wanna say are overwhelmed, but sometimes it’s an additional, I don’t want to say burden either. But where people just feel that ‘I’ve done enough already in my day, and I just can’t put one more thing on my plate’.” [P5]*

Another interviewee discussed how the disproportionate representation of certain communities in the decision-making process could lead to a lack of consideration for the perspectives held by excluded segments of the population:

> *“But when you look at our decision-makers, and those that have the ear of the Council, a lot of them are White men. Sports are predominantly male sports and women’s sports have been pushed to the backburner after the Second World War. [Women] had to fight for anything, any kind of equity within sport. And so the argument that’s used is, well, women’s sports don’t bring in as much revenue, when I would argue that grassroots sports bring in more on an annual basis than the Roughriders do.” [P6]*

### Inseparability of decision-making from the political environment

Interviewees noted that the agendas of decision-makers were inseparable from their broader political environments. For instance, as elected officials, City Councillors often felt that they were beholden to the interests of their constituents. The exact rationale for this mindset varied among decision-makers. Some City Councillors believed that this mindset reflected their duty as elected officials – viewing it as their mandate to advocate on behalf of their constituents, or at least the portions of their constituency that voted for them. For other City Councillors, this mindset seemed to be more of a matter of self-preservation. As elected officials, their continued tenure on the City Council is contingent on the support of their constituents. If they fail to appeal to enough of their constituents, then they run the risk of being voted out at the end of the electoral cycle. In either case, one interviewee recognized that some City Councillors found themselves advocating for positions that they do not necessarily support:

> *“…he [Councillor] has a constituency that’s very conservative. You build a neighbourhood that’s purposely exclusive. He doesn’t agree with it. But he has to represent it, because that’s his mandate. But he gets frustrated, but he’s expressing a genuine position of his constituents.” [P22]*

Furthermore, because City Councillors felt beholden to their constituents, there were instances where City Councillors felt inclined to pass policy proposals that they believed would benefit their respective wards, even if it was to the detriment of the city as a whole. In other words, the accountability that City Councillors felt to their constituents did not necessarily extend to the City of Regina as an entity unto itself. One interviewee made this observation:

> *“You have an elected City Council of 11 people with different views, they represent residents of particular kinds of neighbourhoods that are going to be pressuring them on certain kinds of issues. And sometimes the Official Community Plan is not embraced. Because there’s pushback from residents, they don’t want this to happen, or they want that to happen. And so you’re going to have the elected officials push for that. And it’s not always what’s in the best interest of the city as a whole.” [P25]*

Finally, the agendas of City Councillors were subject to change over time, especially as a reflection of concurrent changes in the prevailing attitudes of their constituents. There are some topics that may garner interest at a certain point in time, only to lose popularity later on. For instance, one interviewee noted how the voting patterns of City Councillors shifted to reflect the waning sympathy of the general public to issues around homelessness:

> *“In June of 2022, they all unanimously passed a motion saying they want to explore what it could mean for the city to go forward and do some more funding. And then in December, almost all of Council was very strong and adamant in their decision around we will not levy more property taxes in support of social services for individuals who are homeless, and their way in doing that, we are going to make a clear stance that it is the provinces role to step in in these issues.” [P4]*

### Relationship with the private sector

Many interviewees recognized that while much of the policy development process occurs in the public sector, the actual implementation of policies often involves actors from the private sector. This can be seen through the crucial role of developers in the implementation of policies through modifications to the built environment. Developers are private entities that are contracted by the municipal government to facilitate changes to the built environment through activities such as: surveying land, renovating properties, installing utilities, and building new infrastructure.

As private entities, developers are not subject to the same level of direct government oversight as those employed directly in the municipal public sector. Furthermore, developers are often driven by profit motivations, meaning they are unlikely to engage with projects that they do not regard as potentially profitable. This can result in developers having different objectives from the City. Even when these parties work towards a common goal – e.g. the development of complete communities that people want to live in – their efforts may be underpinned by different motivations. Developers were often found to be motivated by generating short-to-medium-term profits in order to recoup the costs of development, whereas the City was typically more concerned with ensuring that a given project had the longest lifespan possible.

One interviewee discussed the challenges associated with reconciling their policy-making objectives with the interests of developers:

> *“In Regina, it’s the developers building the neighbourhoods, and the developers have different goals in mind versus the City. We want to have complete communities, sustainable communities, we want to have communities where people want to live in. The developers obviously want to be able to sell the lots. Our goals aren’t necessarily the same.” [P2]*

There are many instances where the profit-oriented motivations of developers are at odds with the objectives of municipal policy- and decision-makers. At the same time, interviewees recognised the importance of developers to the implementation of policies – they are reliant on the cooperation of developers to translate urban planning policies into the real world. Interviewees thus had to ensure that their policy proposals incorporated the motivations of their partners in the private sector as stated by one interviewee:

> *“In terms of implementation, there’s the teeth part of it. And usually that’s a really strong requirement when we’re dealing with third parties, like developers who want to develop something or put something on some land. They’re a very powerful voice, they approach things from the perspective of facilitating the markets delivering what people want. And that additional requirements, as small as they may be, add cost to development, which then gets passed on to the buying public, which has an overall effect of decreasing housing affordability or things like that.” [P27]*

## Discussion

Our interviews with municipal actors engaged with policy- and decision-making around urban design identified a number of barriers to integration of health and equity in urban design policies and decisions.

The lack of a shared, universal understanding and framing of health and equity was identified as a significant barrier to integrating health into urban design and planning. Our findings showed how ideas around health and equity are framed by different municipal stakeholders. To the best of our knowledge this is one of the first studies to explore understanding of municipal actors about health and equity using qualitative methods.^24,34,56^ The power of ideas and how they are framed is of significant importance in policy making because how different actors understand ideas influence policy- and decision making and shape support for initiatives and programs.^24,57–59^ We found that many policy- and decision-makers principally framed health as an individual’s physical state. This interpretation of health falls short of holistic and internationally-recognized definitions of health – for instance, the definition adopted by the WHO in 1948 which describes health as “a state of complete physical, mental and social well-being and not merely the absence of disease and infirmity”.^60^ There is a growing body of literature recognizing the need for a holistic and intersectional framing of health to inform policy- and decision-making. For example, Triguero-Mas et al.^61^ argued that a ‘just ecofeminist approach’ was needed to build healthy cities and meaningfully tackle social inequities. The just ecofeminist approach is founded on a holistic interpretation of health by acknowledging that an individual’s wellness is more than just their physical state and is often influenced by their gender identity and environmental conditions.^61^ This lack of a holistic framing of health, we argue, has influenced the integration of health into urban design policies in our case study. For instance, when asked how health was integrated into their work, most interviewees referred to enablers of physical health, such as cycling infrastructure, hiking trails, and access to healthy foods. This may suggest that non-physical-determinants of health are still undervalued by many policy- and decision-makers within the municipality. For effective policy change towards healthy urban design, it is critical that ideas and discourses around a holistic frame of health become diffused among actors operating within a policy domain.^59^ As such, health to meaningfully integrated into urban planning, it is imperative that City Council not only values the inclusion of health, but also that this inclusion is demanded by residents.

The siloing of institutions within the municipal government was identified as another significant barrier to the meaningful integration of health into policy- and decision-making. Similar findings have been observed in the existing literature. For instance, Williams et al. (2023) not only argued that an intersectional lens should be applied in urban planning for health equity, but also outlined four strategies for accomplishing this: challenging assumptions, fostering cross-sectoral collaboration, co-designing strategies, and looking to established tools and resources.^62^ Also, in a case study of a Norwegian city, Oldeide et al.^63^ identified siloing as one of the biggest hurdles to successfully managing youth drug prevention. Siloing often created confusion regarding the jurisdictional ownership of drug prevention mandates, resulting in uncertainty over which departments are and are not responsible for addressing youth drug prevention.^63^ This was exacerbated by the lack of meaningful opportunities for cross-sectoral collaboration, preventing policy-makers from different departments from effectively pooling together their knowledge and resources.^63^ Lowe et al. (2022) similarly acknowledged the limitations posed by siloing in municipal policy- and decision-making, stating that the creation of healthy and sustainable cities could not occur without integrated urban planning. They argued that this integration must occur both vertically – between different levels of government – and horizontally – between different organizations at the same level of government.^64^ This integration is vital to preventing fractured urban governance and foster the creation of coherent policy frameworks. We argue, the horizontal integration should include the local public health authorities and community organizations, where municipal actors work closely with public health professionals and members of community to more effectively address the SDOH.^65,66^ There is an urgent need to recognize and realize the co-benefits of collaboration between public health and planning workforce.^67^ Giles-Cort et al. (2016) suggest that an integrated urban planning can optimize the use of existing resources by preventing redundant jurisdictional overlap. They argued an integrated planning helps avoid unintended consequences of municipal projects by bringing together the insights of a range of stakeholders with unique perspectives.^1^

Limited legal power of municipalities was identified as another barrier to integration of health and equity in urban design. In Canada, municipal governments have limited autonomy, and are largely recognized to operate within the mandates set out by provincial governments.^68^ The status of municipal governments in Canada differs from that of most American and European municipalities, many of which have constitutionally-enshrined rights and powers that allow them to better resist the actions of higher levels of government.^69^ From a quantitative index developed by Smith & Spicer, it was found that none of Canada’s largest cities have been able to achieve a significant level of fiscal, legal, or political autonomy from their respective provincial governments.^69^ The limited legal power of municipal governments is a point of contention among many municipal policy- and decision-makers.^70^ This relationship has been further strained by the downloading of responsibilities from the provincial government onto municipal governments.^71^ Provincial governments have justified the downloading of responsibilities onto municipal governments under the justification that such measures are intended to increase the autonomy of municipal governments by expanding the scope of their jurisdictions.^72^ In contrast, some municipal governments have countered these assertions by noting that these downloaded responsibilities are often associated with new costs and expenditures.^72^ In many cases, the municipalities themselves are expected to cover the costs of these downloaded responsibilities without receiving a commensurate increase in resource allocation, meaning that policy downloading can effectively result in the limitation of municipal autonomy by stretching their resources thin. ^73^ This precipitous relationship between municipal and provincial governments impedes the effective capacities of municipal policy- and decision-makers.

Finally, it was observed that inadequate representation was another significant barrier to the integration of health particularly equity in urban design policies and decisions. It is important to consider which populations are more likely to be disproportionately represented at the level of municipal government. For example, Heritz (2018) noted that Indigenous peoples are severely underrepresented in Regina’s municipal governance structure, even despite comprising roughly 10% of the city’s population.^74^ Due to their experiences with systemic oppression and marginalization, Indigenous peoples in Canada face ongoing barriers to participating in the institutions that have played a leading role in dismantling their sovereignty, cultural identity, and traditional ways of life.^75^ A similar phenomenon has been observed among immigrants and visible minorities, who tend to be severely underrepresented in the local politics of Canada.^76^ For instance, in Toronto, only 11.1% of City Council seats are held by visible minorities – compared to the 36.8% of the metropolitan population who identify as visible minorities.^76^ As is the case for Indigenous peoples, it has been noted that many of the factors contributing to the political underrepresentation of immigrants and visible minorities have also manifested as structural barriers to their wellbeing – for instance: language barriers, social exclusion, and a lack of familiarity with Canada’s political systems.^64^ Similar factors have been known to contribute to the longstanding underrepresentation of racial or ethnic minorities in municipal governments in the United States.^77^ In many instances, the populations that experience the greatest barriers to engaging in their local governments are the same ones that encounter the greatest health inequities stemming from urban design.

### Study Limitations

One limitation of this study is the type and size of sample we used to collect the data, which may limit the extent to which our findings can be extrapolated. We collected data form the City of Regina, which is a medium-sized city in Western Canada. Although the barriers to integration of health and equity in urban design that we found in this study are specific to the City of Regina, the findings can be generalised to other jurisdictions, especially among cities with similar size in Canada. However, since policies and legislations are inconsistent across different jurisdictions, generalising our findings to the legal or political perspectives may not be acceptable. Therefore, further research is warranted to validate and test the barriers in other jurisdictions and countries.

### Conclusion

This study shed light on some of the major barriers and facilitators to integrating health into the urban design and planning of a medium-sized city in Canada. The findings of this study may be invaluable to urban planners and public health professionals, especially as there has been growing recognition of the potential for cities to play a leading role in enhancing urban population health and health equity. As urbanisation continues to bring a greater share of the world’s population into urban areas, it is imperative that we deepen our understanding of how municipal governance can be leveraged to create environments that are conducive to the wellbeing of their residents.

## Data Availability

All data produced in the present study are available upon reasonable request to the authors.

## Acknowledgements

The authors wish to thank the research participants for their time and insights in the interviews and analytic checks that form the basis of this paper. Dr. Adele Cassola has provided extensive support during the conceptualization phase of this research and reviewing the initial draft of findings. We gratefully acknowledge the collaboration of the City of Regina in recruiting research participants.

